# Post Intensive care outcomes and follow-up in Children: A Collaboration of Health care providers, researchers, and families Utilizing knowledge co-production

**DOI:** 10.1101/2024.10.15.24315556

**Authors:** Michelle Dunphy, Gaby Yang, Jason Marchand, Jenny Retallack

## Abstract

**Background:** Many children do not return to their pre-admission health status following admission to the paediatric intensive care unit (PICU), facing a range of physical, cognitive, emotional, and social challenges collectively known as Post-Intensive Care Syndrome in Paediatrics (PICS-p). The sequelae associated with PICS-p necessitate comprehensive follow-up care intending to address these multifaceted needs. The ***P***ost ***I***ntensive care outcomes and follow-up in ***C***hildren: ***A C***ollaboration of ***H***ealth care providers, researchers, and families ***U***tilizing knowledge co-production (PICACHU) study aims to develop a shared care follow-up service for post-PICU patients and their families. It also seeks to facilitate outcomes research and identify quality improvement (QI) initiatives to mitigate the impact of PICS-p.

**Methods:** The study employs a pragmatic approach informed by the Medical Research Council (MRC) framework and co-design methodology. The research includes surveys and focus group discussions (FGDs) with purposively sampled post-PICU families, acute care pediatricians, community pediatricians, general practitioners (GPs), and primary care nurse practitioners (NPs). Data collection tools include adapted versions of existing surveys and semi-structured interview guides. The analysis will involve qualitative and quantitative methods, utilising SPSS for statistical analysis and NVivo for thematic analysis of FGDs.

**Conclusion:** The PICACHU study is the first of its kind to use a co-design approach to create a post-PICU shared care follow-up service in British Columbia (BC), Canada. The findings will provide valuable insights for improving post-PICU care services in BC and potentially other jurisdictions.

## BACKGROUND

In high-resource countries, advancements in paediatric intensive care have significantly reduced mortality rates (1), yet many survivors face enduring dysfunctions due to their critical illness and the life-saving interventions used during treatment (1-5). This residual burden is known as PICS-p and encompasses four domains: physical, cognitive, emotional, and social impairments (1-3, 5, 6). These impairments can significantly affect the lives of survivors and their families (6-16).

Recognising the need for a broader perspective, Rahmaty et al. (17) extended the original PICS-p framework by incorporating the bioecological theory of human development to reflect children’s interconnectedness within broader social systems. This updated perspective underscores the need for follow-up services that address medical needs and account for socioecological factors like family resources, social connections, and access to specialist healthcare services (17-21).

Clinicians and researchers have increased focus on providing ongoing support to PICU survivors and their families. Despite this focus, significant inconsistencies in post-PICU follow-up care have been identified across Canada (18, 22-26). Many existing services target specific disease processes such as congenital heart disease, extracorporeal support, sepsis, or neurotrauma, leaving the remaining families without structured post-discharge support (26). To address these gaps, the BC Children’s Hospital (BCCH) PICU team, in collaboration with clinicians at Victoria General Hospital (VGH) PICU and Child Health BC (CHBC) are adopting a co-design approach [27, 28] to create a shared care^1^ follow-up service. CHBC is a health improvement network within the Provincial Health Service Authority (PHSA) that focuses on care improvement for health service delivery for children and families across BC.

The co-design approach seeks to ensure that follow-up services are aligned with the needs of survivors and their families. This strategy aims to build follow-up services that account for the complex and intersecting factors that influence children’s recovery and development, addressing the gaps identified in current post-PICU care.

## METHODS AND ANALYSIS

### Aim

The aim of the PICACHU study is to develop a shared care follow-up service to support patients and their families post-PICU admission while facilitating outcomes research and identifying quality improvement (QI) initiatives during admission that could minimise the impact of PICS-p. This project will be approached with an equity lens to ensure that all patients and families, regardless of their background and location within the province, have equal access to care and support.

### Objectives

The objectives of the study are to:

1. Explore the current pathway for general follow-up services required for patients and families after discharge from a PICU in BC.
2. Identify and understand barriers and enablers that acute care paediatricians and community healthcare providers [HCP] (paediatricians, general practitioners, primary care nurse practitioners) encounter in supporting children and families after PICU discharge in BC.
3. Explore the knowledge base of community HCP in BC regarding the sequelae of PICS-p.
4. Identify and explore the domains of PICS-p that children and their families frequently experience within the local context of BC and the challenges encountered after discharge.
5. Actively engage key stakeholders (families, HCP) in the co-design process to develop a follow-up prototype.

## METHOD/DESIGN

### Study Design

The study will be informed by a pragmatic lens using the MRC framework and co-design (summarised in Table 1, Appendix) (28-30) to ensure that the resulting intervention is sustainable, can be replicated across the province, is evidence-informed (28), and produces a service with high user satisfaction (31). The study will unfold in stages, combining research and QI activities to guide the intervention development (Figure 1).

**Table 1.**
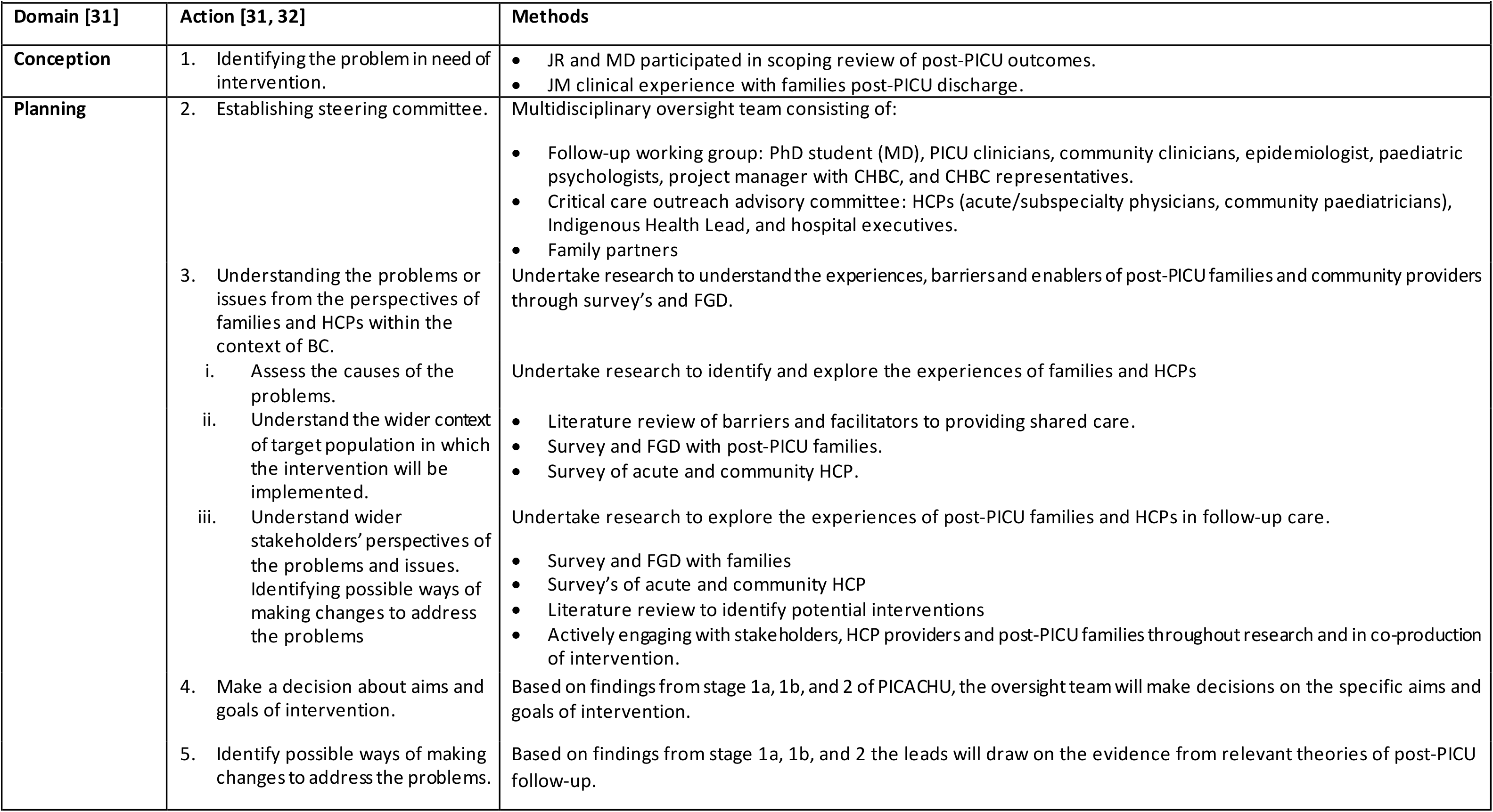
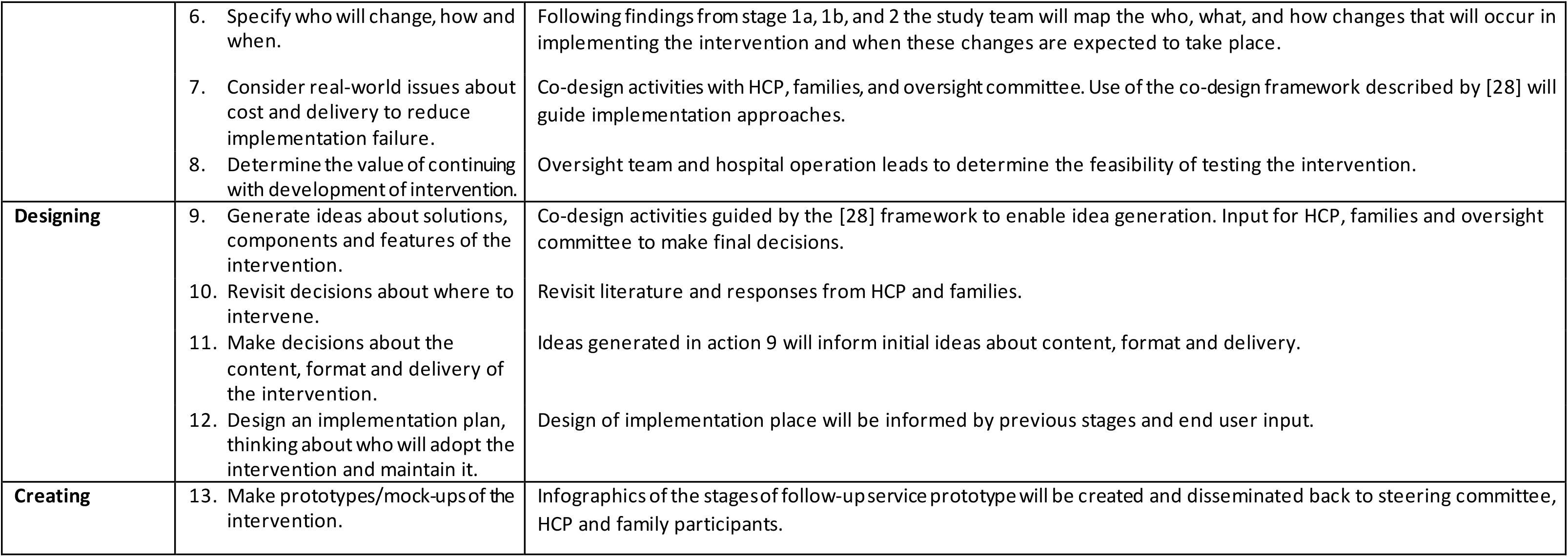
Domains and planned methods to develop a shared care post-PICU follow-up service.

|  |  |  |
| --- | --- | --- |
| Domain [31] | Action [31, 32] | Methods |
| Conception | 1. Identifying the problem in need of intervention. | JR and MD participated in scoping review of post-PICU outcomes.<br>JM clinical experience with families post-PICU discharge. |
| Planning | 2. Establishing steering committee.<br>3. Understanding the problems or issues from the perspectives of families and HCPs within the context of BC.<br>Assess the causes of the problems.<br>Understand the wider context of target population in which the intervention will be implemented.<br>Understand wider stakeholders' perspectives of the problems and issues. Identifying possible ways of making changes to address the problems<br>4. Make a decision about aims and goals of intervention.<br>5. Identify possible ways of making changes to address the problems. | Multidisciplinary oversight team consisting of:<br>Follow-up working group: PhD student (MD), PICU clinicians, community clinicians, epidemiologist, paediatric psychologists, project manager with CHBC, and CHBC representatives.<br>Critical care outreach advisory committee: HCPs (acute/subspecialty physicians, community paediatricians), Indigenous Health Lead, and hospital executives.<br>Family partners<br>Undertake research to understand the experiences, barriers and enablers of post-PICU families and community providers through survey's and FGD.<br>Undertake research to identify and explore the experiences of families and HCPs<br>Literature review of barriers and facilitators to providing shared care.<br>Survey and FGD with post-PICU families.<br>Survey of acute and community HCP.<br>Undertake research to explore the experiences of post-PICU families and HCPs in follow-up care.<br>Survey and FGD with families<br>Survey's of acute and community HCP<br>Literature review to identify potential interventions<br>Actively engaging with stakeholders, HCP providers and post-PICU families throughout research and in co-production of intervention.<br>Based on findings from stage 1a, 1b, and 2 of PICACHU, the oversight team will make decisions on the specific aims and goals of intervention.<br>Based on findings from stage 1a, 1b, and 2 the leads will draw on the evidence from relevant theories of post-PICU follow-up. |
|  | 6. Specify who will change, how and when.<br>7. Consider real-world issues about cost and delivery to reduce implementation failure.<br>8. Determine the value of continuing with development of intervention. | Following findings from stage 1a, 1b, and 2 the study team will map the who, what, and how changes that will occur in implementing the intervention and when these changes are expected to take place.<br>Co-design activities with HCP, families, and oversight committee. Use of the co-design framework described by [28] will guide implementation approaches.<br>Oversight team and hospital operation leads to determine the feasibility of testing the intervention. |
| **Designing** | 9. Generate ideas about solutions, components and features of the intervention.<br>10. Revisit decisions about where to intervene.<br>11. Make decisions about the content, format and delivery of the intervention.<br>12. Design an implementation plan, thinking about who will adopt the intervention and maintain it. | Co-design activities guided by the [28] framework to enable idea generation. Input for HCP, families and oversight committee to make final decisions.<br>Revisit literature and responses from HCP and families.<br>Ideas generated in action 9 will inform initial ideas about content, format and delivery.<br>Design of implementation place will be informed by previous stages and end user input. |
| **Creating** | 13. Make prototypes/mock-ups of the intervention. | Infographics of the stages of follow-up service prototype will be created and disseminated back to steering committee, HCP and family participants. |

**Figure 1.**
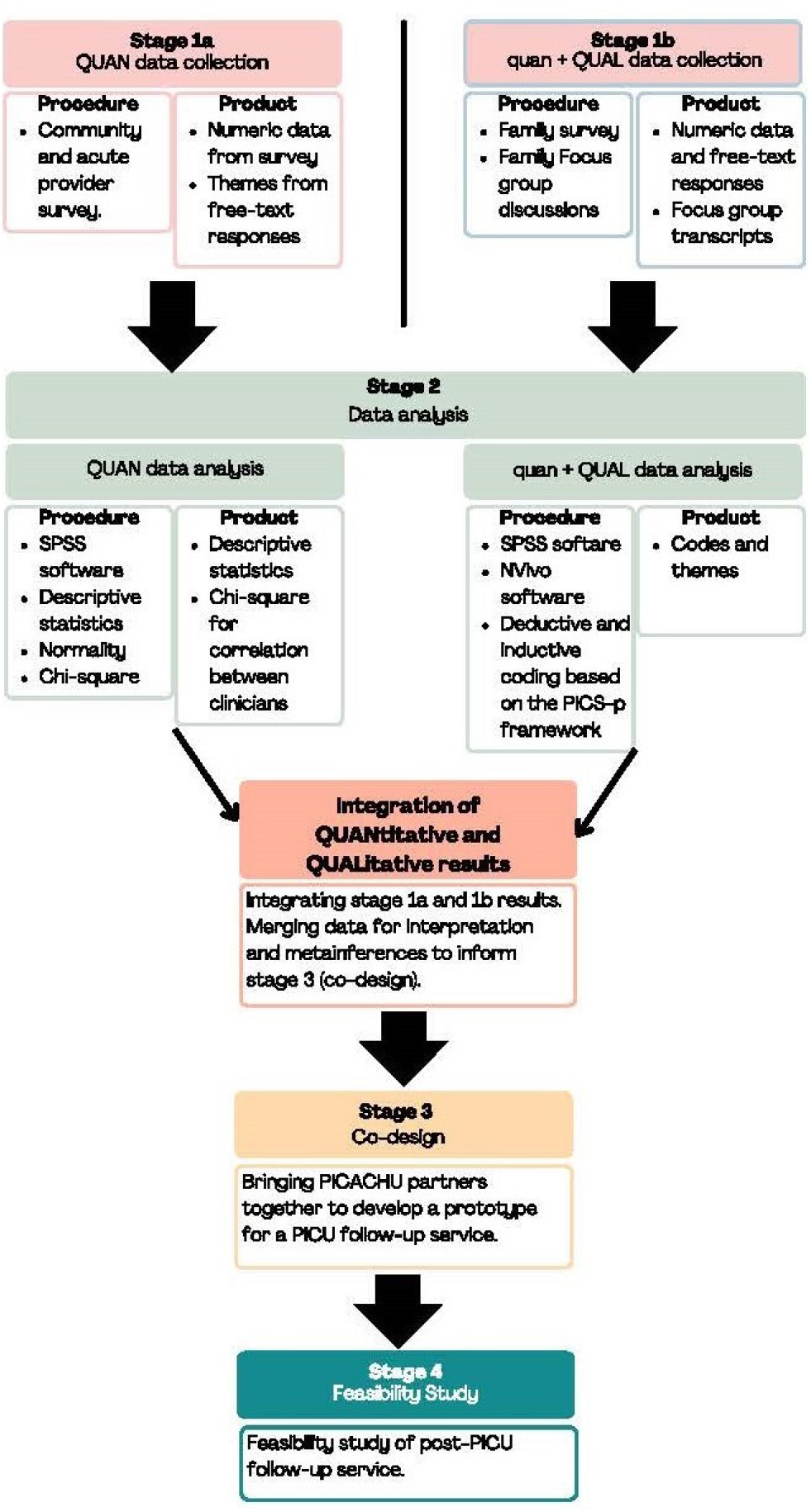
Stages of the PICACHU study (QUAN = quantitative; QUAL = qualitative).

### Participants

Purposive sampling of post-PICU families, acute care paediatricians (BCCH and VGH), community HCP (paediatricians, General Practitioners [GP] and primary care Nurse Practitioners [NP]).

### Sample and Recruitment

#### Stage 1a: Healthcare Provider survey

Informal communication channels (closed-secure health authority email system) will be used to engage acute care paediatricians. Multiple recruitment strategies will be used to engage with community HCPs, including community paediatricians, GPs, and NPs. An infographic advertising the survey will be distributed using multiple channels, including social media sites (LinkedIn, closed Facebook groups), health authorities and professional registration body’s media channels (Family Doctors of BC, BC Pediatric Society, and Nurse Practitioners of BC) and CHBC networks. Community providers must have experience treating paediatric patients (i.e., adult-only clinicians will be excluded from participating). The survey will only allow clinicians treating paediatric patients to complete it. At the end of stage 1a, providers will be invited to participate on co-designing a shared care model. Community HCPs will be asked to provide contact details for the co-design process.

#### Stage 1b: family consultation

The Virtual Pediatric System (VPS) will be reviewed by the research nurse (MD) to identify families of patients admitted to BCCH PICU who meet the inclusion criteria (Table 2). Island Health employee (GY) will manually review patients at VGH PICU who meet inclusion criteria. If the initial timeframe does not yield a sufficient sample size (at least 60 patients), the timeframe will be expanded to <8 months post-discharge of the project start date. The target sample size will be 15 to 30 family representatives.

**Table 2.** Family participant inclusion and exclusion criteria

| Inclusion criteria | Exclusion criteria |
| --- | --- |
| Families of a child aged 0 to <17 years and discharged from PICU > 1 month but <6 months of project start date. | Patients currently receiving follow-up services<sup>2</sup>. |
| Admission to PICU for ≥72 hours | Transferred to an acute rehabilitation facility |
| Required support for organ dysfunction |  |

Families meeting inclusion criteria will receive a letter notifying them of the study, with the option of “opting out” of the study as per PHSA’s organisational policy. MD will contact families using a telephone consent script to receive further information regarding stage 1b of the project (Figure 1). Written consent will be sent via REDCap electronic data capture tools hosted at BCCH Research Institute, which will include the project outline, reimbursement details, and a poll to solicit participants’ availabilities for the FGD. Once respondents have identified their availability for FGD,

MD will allocate them into groups of 3-5 participants. An email confirmation of the date for their FGD and relevant contact information will be sent out via REDCap. Surveys will be distributed via REDCap one week before FGD. 48 hours before FGD, MD will telephone families to confirm attendance and financial reimbursement details and address family questions.

At the end of FGD (stage 1b), family participants will be given the opportunity to participate in the co-design process. Due to the time difference between stage 1b and stage 3, families will be reconsented at the start of the co-design process.

### Data Collection

#### Stage 1a: community HCP survey

The survey will use an adapted version of the “General practitioner perspectives on a shared-care model for paediatric patients post-intensive care: A cross-sectional survey” (32). It will be distributed using the same channels as recruitment. Participants will be directed to the online survey in REDCap. The survey will be open for responses over a four-week period.

#### Stage 1a: acute care paediatricians survey

The survey will contain six questions, one of which invites the provider to participate on the co-design of the follow-up service. The survey will be sent via the closed health authority email system. Responses will be inputted into a password-protected Excel spreadsheet.

#### Stage 1b: family consultation

Family consultation will involve a survey and FGD. The survey will contain demographic information about the child and family, length of admission, and services support accessed post-discharge, as shown in Additional file 2. It will be disseminated in English electronically via REDCap.

The research team will conduct the FGD, which will be recorded using the University of British Columbia (UBC) Zoom account (JR). The FGD facilitators will use an open-ended, semi-structured interview guide to elicit a narrative from the parents/caregivers’ experiences following discharge from the hospital. They may also prompt or probe participants’ responses.

### Data Analysis

#### Stage 1a: Acute care and community HCP survey responses

Data will be imported into IBM SPSS Statistics for Windows™ version 29 (IBM Corp: Armonk, NY) from REDCap (community HCP survey) or a password-protected Excel™ Spreadsheet (acute care paediatrician survey).

Analysis related to the specific objectives of the project include:

*Objective 1: Explore the current pathway for general follow-up services required for patients and families after discharge from a PICU in BC*.

*Objective 2: Identify and understand barriers and enablers that acute care paediatricans and community HCP encounter in supporting children and families after PICU discharge in BC*.

*Objective 3: Explore the knowledge base of community HCP in BC regarding the sequelae of PICS-p*.

Descriptive statistics will be presented for community HCP demographic information (e.g., paediatrician, GP, NP) and frequency distributions of the responses. Chi-square testing will be used to determine a correlation between paediatricians and primary HCPs (GPs and NPs). MD and JR will examine free text responses for common themes. Themes related to barriers and enablers will be mapped using the logic model.

#### Stage 1b: family consultation

*Objective 4: Identify and explore the domains of PICS-p that children and their families frequently experience within the local context of BC, and the challenges encountered after discharge*.

Survey data will be inputted directly into the REDCap database and imported into IBM SPSS Statistics for Windows™ version 29 (IBM Corp: Armok, NY). Data will be summarised using means and medians for continuous data and proportions for categorical data. MD will thematically analyse open-ended free-text responses.

FGD will be audio recorded using UBC Zoom transcription services. MD and JR will read the transcriptions in their entirety to ensure accuracy of the transcription. MD and JM will analyse and code FGD data. Qualitative description will inform the initial analysis, allowing for a rich and nuanced understanding of the family’s challenges (33, 34). The coding and categorising of the data will occur via QSR International’s Nvivo software (14). Member checking will be used to ensure the rigour and validity of the findings.

#### Stage 2: collating findings and drawing metainferences

MD and JR will analyse the findings at the end of stages one and two using a concurrent transformative mixed methods approach (Figure 1). The oversight team (Table 1) will then consider common themes, barriers, enablers, and end users’ experiences. The information obtained from stages 2 will inform the co-design activities in stage 3.

#### Stage 3: co-design process

The team will follow a sequential co-design approach integrating evidence from published literature, expert knowledge, and end-user involvement in designing the initial prototype, (29, 35) involving a range of methods to ensure the perspectives of end-users and HCPs are captured. Due to the geographic diversity of the province, engagement will be predominantly virtual (e.g., REDCap survey or secure virtual meeting platforms such as Zoom). Acknowledging the time demands on clinicians and families, co-design activities will not exceed one hour. At the end of each design activity, final decisions will be communicated back to participants to ensure closed-loop communication.

### ETHICS

#### Stage 1a: community provider survey

This project stage is considered quality improvement (QI) and will not require approval from the research ethics board. It will be reviewed by the PHSA research, quality improvement, and privacy board before distribution.

#### Stage 1b: family consultation

Submission for approval from the institutional research ethics board (UBC RISe) was completed (H23-01929) before the commencement of research activities within the project. Participants will be informed of the voluntary nature of the project and consent, which can be withdrawn at any time. After completing the consent form, parents/caregivers will be financially reimbursed for their time ($25 per hour) cheque payments. There are no physical risks associated with this study. However, there is a potential risk that families may experience distress and/or be re-traumatised by discussing their experiences. Options to access resources for support will be provided to the parents/caregivers in the survey consent and before commencing the FGD sessions. Responses will be de-identified for analysis, and any shared findings will be de-identified.

#### Zoom considerations

The ethical considerations of using UBC Zoom will be outlined in the consent documents to participants. Before recording the session, these will be reiterated at the start of each FGD.

#### Provisions for Consent

Due to the nature of the project and in consultation with the Indigenous Research & Evaluation Specialist at PHSA, families will be offered the opportunity to have an Elder to support the FGD. In addition, the use of colonising research language will be changed in consent documents to “sharing circle” from FGD. During initial contact with families, whether they identify as a First Nation family will be unknown. In recognition of the deleterious impact of colonising research practices on First Nations People of Canada, MD will offer all families the opportunity to have the sharing circle (FGD) supported by an Elder. Honorariums will be provided to Indigenous Elders from the project funds.

The family survey has not been translated from English. At least one family member must speak sufficient English to provide consent. Statistics Canada (36) states that 95% of B.C. residents list English as their first language. Completing the survey is not mandatory to attend the FGD, and families requiring a translator for the FGD session will be facilitated if they wish to attend. In this situation, verbal consent will be obtained via an interpreter. During the FGD, families will be offered the opportunity to complete the survey, which will be supported by interpreter services.

### Patient and Public Involvement

Feedback on the study design and materials (consent, survey, FGD questions) was reviewed and amended based on the feedback of the family partners.

## KNOWLEDGE DISSEMINATION

Findings from the PICACHU study and follow-up prototype will be disseminated via a provincial newsletter. The project team will be guided by the expertise of the Indigenous Research & Evaluation Specialists at PHSA on how to proceed with the de-identified findings from Elder-supported FGDs to prevent the appropriation of First Nation families’ knowledge.

Beyond the local BC context, a multifaceted strategywill be employed to widely share the results, including professional journals and high-quality, peer-reviewed, open-access journals. A summary of findings will also be distributed to the Canadian post-PICU Follow-up collaborative. Given the emerging nature of this field of study, the results could impact the delivery and configuration of follow-up services in other jurisdictions, potentially improving services for post-PICU patients and their families.

## DISCUSSION

To our knowledge, the PICACHU study will be the first co-design of a post-PICU shared care follow-up service using a pragmatic mixed-methods approach. The project design supports a pragmatic philosophy with a real-world application, allowing the project team to understand the phenomena within the context of BC and will provide valuable insights to inform decision-making and future practice.

Due to space limitations, the final stage (stage 4) of the PICACHU study has not been discussed in this protocol and will be presented in a separate publication. Stage 4 will entail a feasibility study of the prototype developed from stages 1 to 3.

### Limitations

We acknowledge no child representatives will be participating in the co-design stage of the

PICACHU study. Stage 4 of the PICACHU study will incorporate the experiences of child survivors of critical illness.

## Data Availability

Data can be requested from 1 year after study completion from the principal investigator (JR).

## DECLARATIONS

## Acknowledgements

The authors would like to acknowledge this work was primarily carried out on the unceded land of the Lekwungen, x^w^məθkwəyəm (Musqueam), Skwxwú7mesh Úxwumixw (Squamish), and səlililwətaʔɬ (Tsleil-Waututh) Nations. The authors would like to acknowledge the following: Lori Anthony, MClinEpi, Amie Hilder, BPhysio, Dana Newcomb, MBBS FRACGP, Kerri-Lyn Webb, MBBS, James Best, MBBS FRACGP, Christian Stocker, MBBS MD FMH (CH), Debbie Long, RN PhD for permission to use the survey from their study “General practitioner perspectives on a shared-care model for paediatric patients post-intensive care: A cross-sectional survey.” We thank Meghan O’Neill (Child Health BC) for creating REDCap survey instruments and Dr Srin Murthy (BC Children’s Hospital, PICU Intensivist) for consulting on the article.

## Competing Interests

The following authors declare that they have no competing interests: MD, JR, GY, JM.

### Authors’ Contributions

JR and MD designed the study in consultation with GY and the CHBC steering committee. JR wrote the questions for the FGD and family surveys. MD amended the community provider survey. MD and JR wrote the acute provider survey questions. MD wrote the article, which was reviewed by JR, GY, and JM. JM assisted in the data analysis of FGD. All authors reviewed the article for important intellectual content. All authors read and approved the final manuscript.

### Funding

The project is funded by the BC Children’s Hospital Foundation (BCCHF), which is not involved in the study design, data collection, analysis, and interpretation. Additionally, BCCHF does not provide input in the article’s writing or the decision to submit the results for publication. BC Children’s Hospital Foundation provides open-access funding.

### Availability of Data and Materials

Data can be requested from 1 year after study completion from the principal investigator (JR).

### Patient and Public Involvement

Patients and/or the public were involved in this research’s design, conduct, reporting, or dissemination plans. For further details, refer to the Methods section.

### Consent for Publication

All authors carefully read the manuscript and approved it for publication.

## Footnotes

1 The definition of a shared care model is based on the description by Vuong et al. (2020), which sees care as a coordinated approach that brings together providers from different specialties, including primary and community care, to collaborate on a person’s care for as long as needed.

2 The following patients currently receive follow-up services from their primary treating team within BC: Technology-dependent children (e.g., home tracheostomy and ventilation program), oncology patients, non-accidental injury, solid organ transplant, and bereaved families.

